# Contact with young children is a major risk factor for pneumococcal colonization in older adults

**DOI:** 10.1101/2024.01.03.24300789

**Authors:** Anne L. Wyllie, Devyn Yolda-Carr, Maikel S. Hislop, Sidiya Mbodj, Loren Wurst, Pari Waghela, Ronika Alexander-Parrish, Lindsay R. Grant, Adriano Arguedas, Bradford D. Gessner, Daniel M. Weinberger

**Author notes:** These authors contributed equally to the study (Anne L. Wyllie, Daniel M. Weinberger). **Corresponding author:** Anne Wyllie, PhD, Yale School of Public Health, LEPH 823, 60 College St, New Haven, CT 06510, USA.

## Abstract

**Background:** Important questions remain about the sources of transmission of pneumococcus to older adults in the community. This is a critical question for understanding the potential indirect effects of using pneumococcal conjugate vaccines (PCVs) in children and older adults. For non-institutionalized individuals, the most likely source of adult-to-adult transmission is in the household. The goal of this study was to characterize the dynamics and risk factors for acquisition of pneumococcus in older adults.

**Methods:** We designed a longitudinal study to sample adults >60 years of age living in the same household (New Haven, CT, USA), and without younger contacts residing in the household. Saliva samples and questionnaires regarding social behaviors and health status were obtained every 2 weeks for a period of 10 weeks. DNA extracted from culture-enriched saliva was tested using qPCR for pneumococcus genes *piaB* and *lytA*.

**Results:** Across two study seasons (November 2020-August 2021, November 2021-September 2022), 121 individuals from 61 households were followed for 6 study visits; 62 individuals were enrolled in both seasons. Overall, 52/1088 (4.8%) samples tested positive for pneumococcus based on *piaB*, with 27/121 (22.3%) individuals colonized on at least one time point. Several individuals were colonized at multiple timepoints including two individuals who were colonized throughout the 10-week sampling period; two others were colonized at 5 of 6 time points. In 5 instances, both members of the household were carriers in the same season, though not necessarily at the same time point. Pneumococcal carriage was substantially higher among individuals who had contact with children (10.0% vs 1.6%). Participants who reported recent contact with <5-year-olds and 5-9-year-olds had particularly elevated prevalence (13.8%; 14.1%, respectively).

**Conclusions:** Contact with young children was the most important factor that influenced pneumococcal acquisition rates. While there were several instances where both adult household members were colonized at the same time or at sequential visits, these individuals also both typically had contact with children.

## INTRODUCTION

*Streptococcus pneumoniae* (pneumococcus) is an important human commensal and cause of respiratory infection and invasive disease. Pneumococcus resides primarily in the upper respiratory tract and is transmitted via aerosolized droplets. Studies frequently report children as the main reservoir of transmission of pneumococcus in the community.^1–6^ It is clear that vaccinating infants with pneumococcal conjugate vaccines (PCVs) leads to sharp reductions in disease caused by vaccine-targeted serotypes in unvaccinated age groups. However, some PCV serotypes (such as 3 and 19A) have persisted as causes of disease in the adult population,^7^ despite PCVs being used at high rates among infants. This has led to questions about whether these serotypes might be transmitted by age groups other than children.

If substantial pneumococcal transmission occurs between adults, then vaccination of older adults could have an additional benefit of reducing transmission and subsequent disease. However, quantifying carriage among adults is challenging. Surveys of pneumococcal colonization in older adults have reported vastly different estimates, depending on the sampling and testing methodology used. Using culture-based detection of pneumococcus from nasopharyngeal swabs,^8^ carriage is rarely detected (<5%). When saliva samples are tested using qPCR, higher rates of colonization are detected. Regardless of the method used, no study has directly evaluated rates of transmission among community-dwelling adults, with prior studies focused on households where either children or adults could be the source of transmission.^9–12^

The risk of adult pneumococcal colonization is likely influenced by numerous factors including contact patterns, living conditions, and underlying individual health. In this study, we aimed to evaluate the importance of within-household transmission between adults, and risks associated with pneumococcal acquisition among community dwelling older adults.

## METHODS

### Ethics

This study was approved by the Institutional Review Board at Yale School of Medicine (Protocol ID #2000026100). Demographic data and samples were collected after the study participant had acknowledged that they had understood the study protocol and provided digitally signed-informed consent, collected in the Research Electronic Data Capture (REDCap) electronic data capture tools hosted at Yale University.^13,14^ All participant information and samples collected were assigned anonymized study identifiers.

### Study design

Using a longitudinal household sampling design, household pairs (e.g. married couples) were enrolled if both individuals were age 60 or above and no additional individuals under the age of 60 years were residing in the household. If an individual reported symptoms of respiratory illness at time of consenting or had received antibiotics or pneumococcal vaccination within the past four weeks, the enrollment of that household pair into the study was delayed by up to four weeks. There were no exclusion criteria based on underlying health status. Enrollment was completed over the course of the two autumn/winter seasons of 2020/2021 and 2021/2022. Saliva samples and demographic data were collected every 2 weeks for 6 total visits (10 week study inclusion), as previously described.^6^ At the final visit, study participants also provided a urine sample. Participants from the first season were invited to participate again in the second season.

### Sample processing and pneumococcal detection

On arrival at the lab, raw saliva samples were culture-enriched on TSAII plates with 5% sheep’s blood and 10% gentamicin, as previously described.^6^ DNA was extracted from 200 _μ_l of each culture-enriched sample using a modified protocol of the MagMAX Viral/Pathogen Nucleic Acid Isolation kit (ThermoFisher Scientific) on the KingFisher Apex (ThermoFisher Scientific).^6^ Extracted DNA was tested by qPCR on a CFX96 Touch (Bio-Rad) using primers and probes specific for two pneumococcal genes: *piaB*^15^ and *lytA,*^16^ as previously described^6^. Since the *lytA* gene is not specific to pneumococcus, with homologues present in many other alpha-hemolitic-*Streptococci,*^17^ a sample was only classified as positive for pneumococcus when reporting a *piaB* cycle threshold (Ct) value of <40.^6^ Samples which reported a Ct value 35-40 were tested again and if discrepant (Ct>40), we repeated a third time as a tiebreaker to determine positivity.^6^ All samples were also tested for SP2020^18^ which has been proposed as an additional target to further increase the specificity of detection of pneumococcus.

DNA samples were subsequently pooled, then tested in each of seven multiplexed PCR assays targeting a total of 36 pneumococcal serotypes (see Supplementary Information).^19–22^ In season one, samples testing negative for pia*B* were pooled by 10. Samples that also tested negative for *lytA* were pooled separately from those that tested positive for *lytA.* In both seasons, DNA samples testing *piaB-*positive were pooled by four. Where possible, samples from the same person were pooled together. From each pool, 8 µl of DNA was tested in a total PCR reaction volume of 25 µl, containing 12.5 µl NEB Luna Mastermix and 4.5 µl of primer/probes (see Supplementary Information). All samples from any pool generating a serotype-specific Ct value <40 were re-tested individually in that serotyping assay. The serotype of an individual sample was assessed based on the concordance between the *piaB* Ct value and the serotype-specific Ct value. Testing of negative samples in season one was conducted to evaluate rates of confounding (a serotype-specific Ct value of <40, despite DNA templates being qPCR-negative for *piaB*) within each assay and to inform its reliability.

### Strain isolation

Saliva samples that tested positive for *piaB* with a Ct <28 were re-visited by culture^6^ and/or magnetic bead-based separation (MBS)^23^ in an attempt to isolate pure pneumococci. Pneumococcal isolates were serotyped by latex agglutination and qPCR then stored at −80°C.

### Detection of respiratory viruses

All saliva samples were also tested for the presence of SARS-CoV-2, influenza A/B, and respiratory syncytial virus (RSV). Lysates were prepared from 50 µl of each saliva sample which was heated at 95°C for 5 minutes^24^ before testing in a modified “SalivaDirect” PCR assay,^25^ expanded for multiplexed detection of these viruses.^26^

### Detection of pneumococcus using urine antigen detection (UAD)

On arrival at the lab, urine samples were aliquoted into PIPES buffer. Aliquots were stored frozen at −80°C until batch shipping on dry ice to the reference laboratory of Pfizer Vaccine Research (Pearl River, NY). Upon receipt, samples were stored at −80°C until batched testing could be performed. All samples were tested according to the manufacturer’s protocol using the serotype-specific UAD assays which targets 24 of the 100 known pneumococcal serotypes^27,28^ and the BinaxNOW® test which targets a pan-pneumococcal antigen to determine the presence of any pneumococcus of serotype not covered by the UAD.

### Statistical analysis

Differences in the frequency of categorical outcomes were compared using Fisher’s Exact test in the R Statistical Software v4.1.2.^29^ Factors associated with pneumococcal carriage were evaluated with a multivariate log-binomial model. This was fit using generalized estimating equations (GEE) to account for repeated measures within individuals. Observations for individuals who enrolled in both study seasons were considered a single unit for the GEE (up to 12 observations per individual). An exchangeable correlation structure was used.

Longitudinal analyses of carriage acquisition and clearance were performed using a continuous time Markov Model using the msm package^30^ in R. The model had 2 states (colonized and uncolonized), and we evaluated factors associated with acquisition (transition from uncolonized to colonized) and clearance (transition from colonized to uncolonized). Due to the gap in sampling between seasons and the short mean duration of colonization, data from individuals who were re-enrolled in seasons 1 and 2 were treated as distinct participants. Because we were not able to determine serotype for many carriers, and because there were not clear instances where an individual switched serotypes or had different serotypes within the household, we tracked pneumococcal colonization status but did not account for serotype in this analysis.

## RESULTS

### Population characteristics

From November 2020 through September 2022, 121 individuals from 61 households were sampled; 62 individuals were enrolled in both seasons. In season one (S1), individuals were sampled between November 2020 and August 2021. In season two (S2), individuals were sampled between November 2021 and September 2022. One household was composed of a single individual who was enrolled into the study due to residing in a living facility for older adults.

The mean age of study participants was 70.9 years (range 60-86) (Supplementary Figure 1). Of the study participants, 85.2% were white (Table 1). Among the study participants who responded to the question, 77.6% held a bachelor’s degree or higher. After receiving a high rate of survey incompleteness during season one, we updated our data capture to ensure more complete data in season two.

**Table 1:** Characteristics of the study population.

| Table 1: Characteristics of the study population |  |
| --- | --- |
|  | (N=121) |
| **Age (years)** |  |
| Mean (SD) | 70.9 (5.98) |
| Median [Min, Max] | 71.0 [60.0, 86.0] |
| Missing | 1 (0.8%) |
| **Gender** |  |
| F | 62 (51.2%) |
| M | 59 (48.8%) |
| **Race** |  |
| Asian | 3 (2.5%) |
| Black or African American | 3 (2.5%) |
| White | 105 (86.8%) |
| Missing | 10 (8.3%) |
| **Ethnicity** |  |
| Hispanic or Latino | 1 (0.8%) |
| non-Hispanic | 116 (95.9%) |
| Missing | 4 (3.3%) |
| **Ever smoke?** |  |
| No | 83 (68.6%) |
| Yes | 27 (22.3%) |
| Missing | 11 (9.1%) |
| **Diabetes** |  |
| No | 101 (83.5%) |
| Yes | 8 (6.6%) |
| Missing | 12 (9.9%) |
| **Asthma** |  |
| No | 100 (82.6%) |
| Yes | 5 (4.1%) |
| Missing | 16 (13.2%) |
| **Influenza Vaccine?** |  |
| No | 4 (3.3%) |
| Yes | 103 (85.1%) |
| Missing | 14 (11.6%) |
| **Education level** |  |
| High School | 11 (9.1%) |
| Undergraduate (Bachelor or Associate) | 44 (36.4%) |
| Graduate | 50 (41.3%) |
| Unknown | 16 (13.2%) |

Of the 1,091 samples collected, three were not tested due to low collection volume (n=1) or a weather-related delay (two weeks) in transporting samples to the lab (n=2). Four samples from one individual were not collected because that participant was hospitalized. Three samples from three individuals were not obtained because the participant was unable to produce saliva at that sampling moment.

In 2020/2021, all individuals tested negative for SARS-CoV-2. In 2021/2022, 12 individuals tested positive, one of which also tested positive for pneumococcus at the same sampling moment (#61, serotype 22F/A). None of the samples tested positive for influenza or RSV.

### Prevalence of pneumococcal carriage

Overall, 52/1088 (4.8%) samples tested positive for pneumococcus based on *piaB*, with 27/121 (22.3%) individuals colonized on at least one time point. By study season, 14/95 (14.7%) individuals were colonized on at least one time point in season 1 and 14/88 (15.9%) individuals were colonized at least once in season 2 (Figure 1). In 2/48 (4.2%) households in season 1 and 3/44 (6.8%) households in season 2, both members were carriers, though not necessarily at the same time point. There were not meaningful differences in prevalence across seasons or both demographics (Table S1).

**Figure 1.**
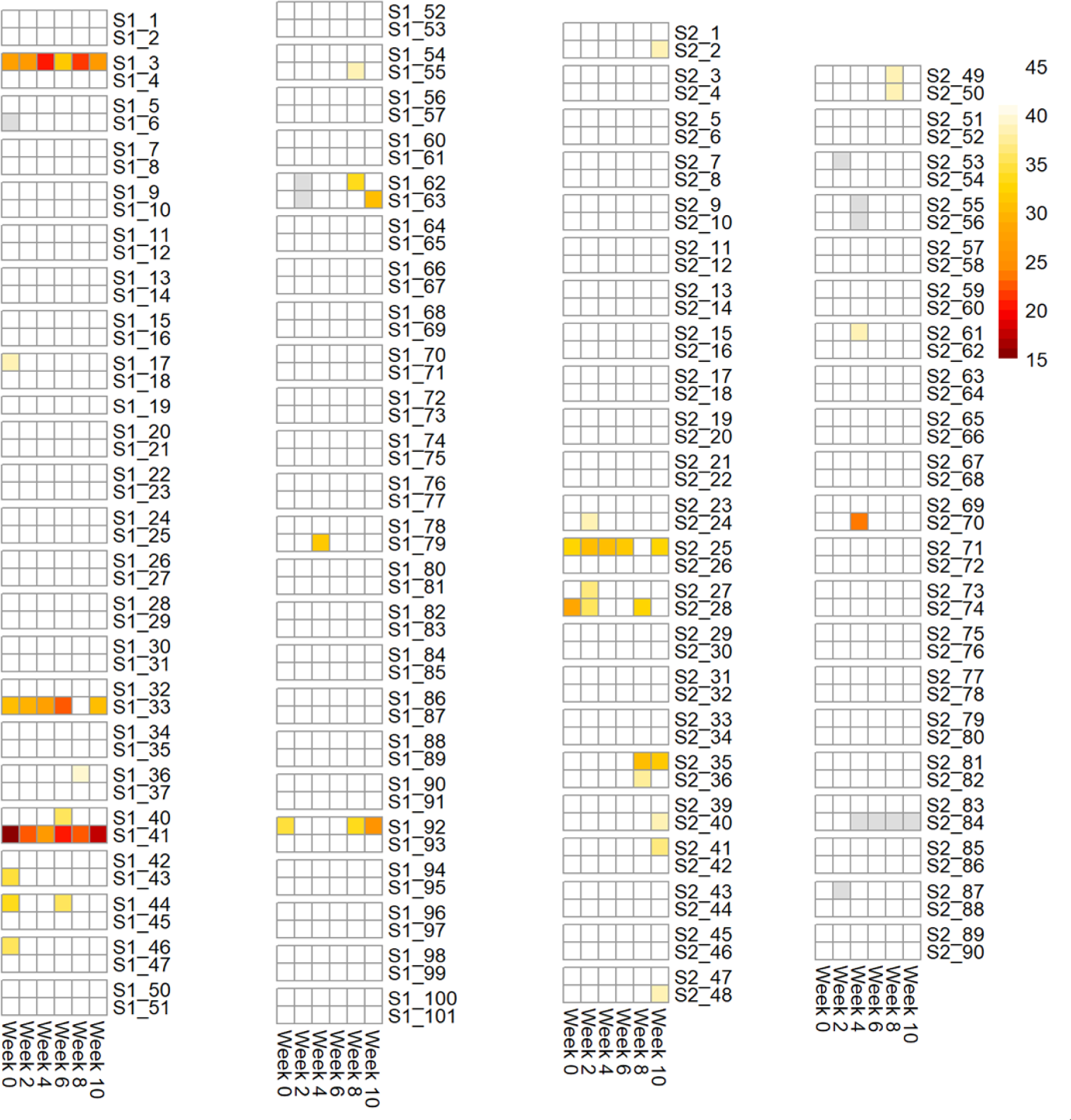
Heatmap of sample positivity for pneumococcus gene, *piaB*. Overall, 52/1088 (4.8%) samples tested qPCR-positive, with 28/183 (15.3%) individuals colonized on at least one time point. Darker colors indicate lower Ct values (higher concentration of pneumococcus). Gray boxes indicate the sample was not tested or missing. Each row represents an individual, each column a time point. Individuals in the same household are grouped together. For participants that were re-enrolled, the study ID does not match between season 1 and 2.

When samples were positive for both *piaB* and *lytA*, there was good concordance in the bacterial density (Ct value) (Figure 2). This was also observed when testing samples for SP2020 (Supplementary Figure 2). However, concordance between *lytA* and SP2020 was weaker, with high rates of samples positive for only one of these targets, indicating a reduced specificity of both the *lytA*^31^ and SP2020 qPCR assays (Supplementary Figure 3) and as such, supported our reasoning to rely solely on *piaB* for determining sample positivity for pneumococcus. As expected for these carriers without pneumonia, none of the urine samples collected tested positive for pneumococcal antigens using either the UAD or the BinaxNOW® test.

**Figure 2.**
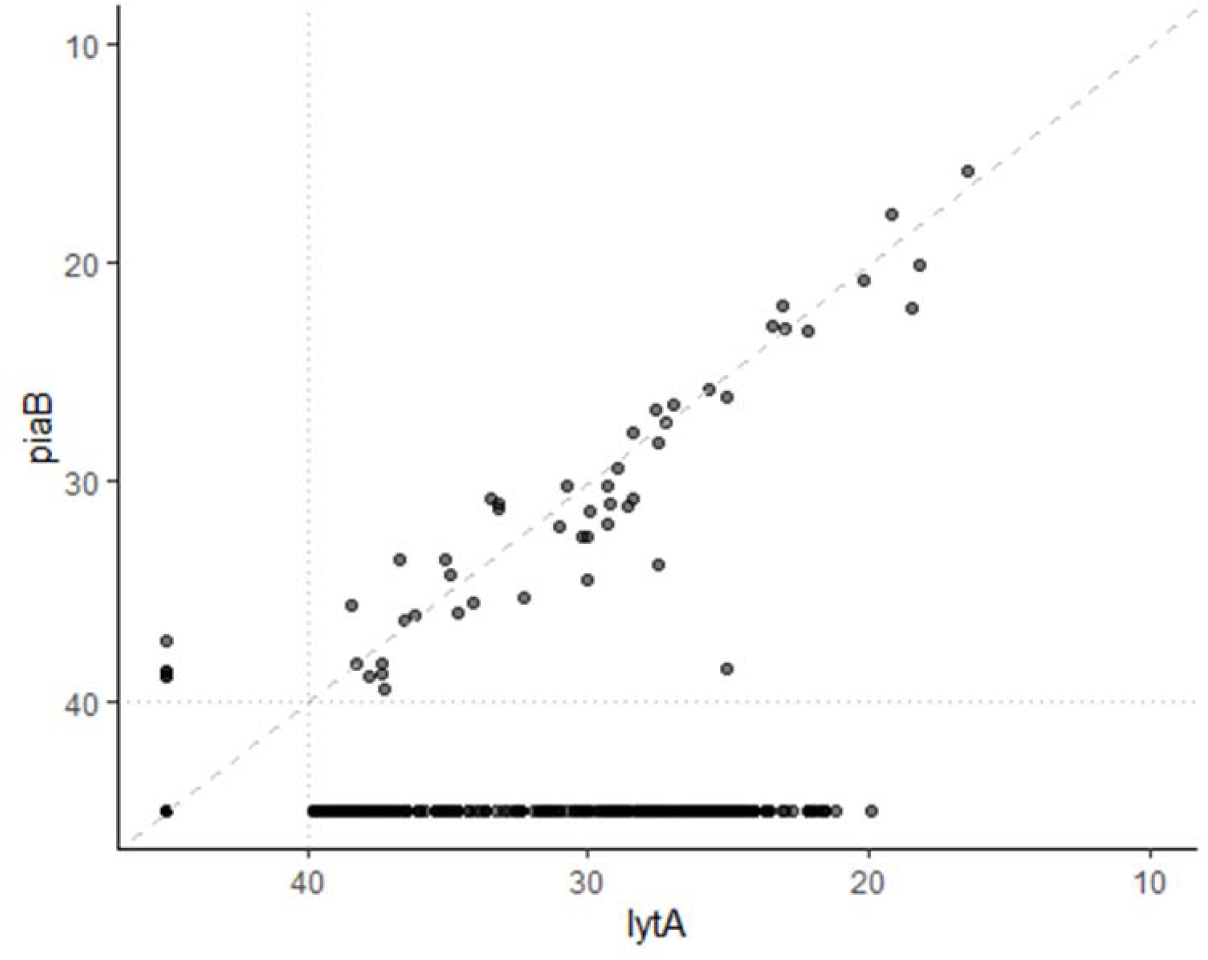
Concordance between *lytA* and *piaB* qPCR values. Ct values are shown, with a value of <40 considered positive for the gene target.

### Pneumococcal carriage and serotype patterns across study seasons

Several individuals were colonized at multiple timepoints including two individuals who were colonized throughout the 10 weeks sampling period in one season (S1 #3, serotype 17F and S1 #41, serotype 15B/C; Figure 3). The household pair of S1 #44 (S1 #45) also tested qPCR-positive for 15B/C at one of the same sampling moments. Two individuals were colonized at five of the six time points (S1 #33, serotype 15B/C and S2 #25, serogroup 6). Participant S2 #25 also participated in the first study season and was positive for pneumococcus on 3/6 sampling moments (S1 #92), with the later two visits of season one also being positive for serogroup 6 (we were unable to resolve a serotype for the first sampling moment they tested positive). This individual reported daily contact with children 2-59 months and 5-9 years of age in both seasons.

**Figure 3.**
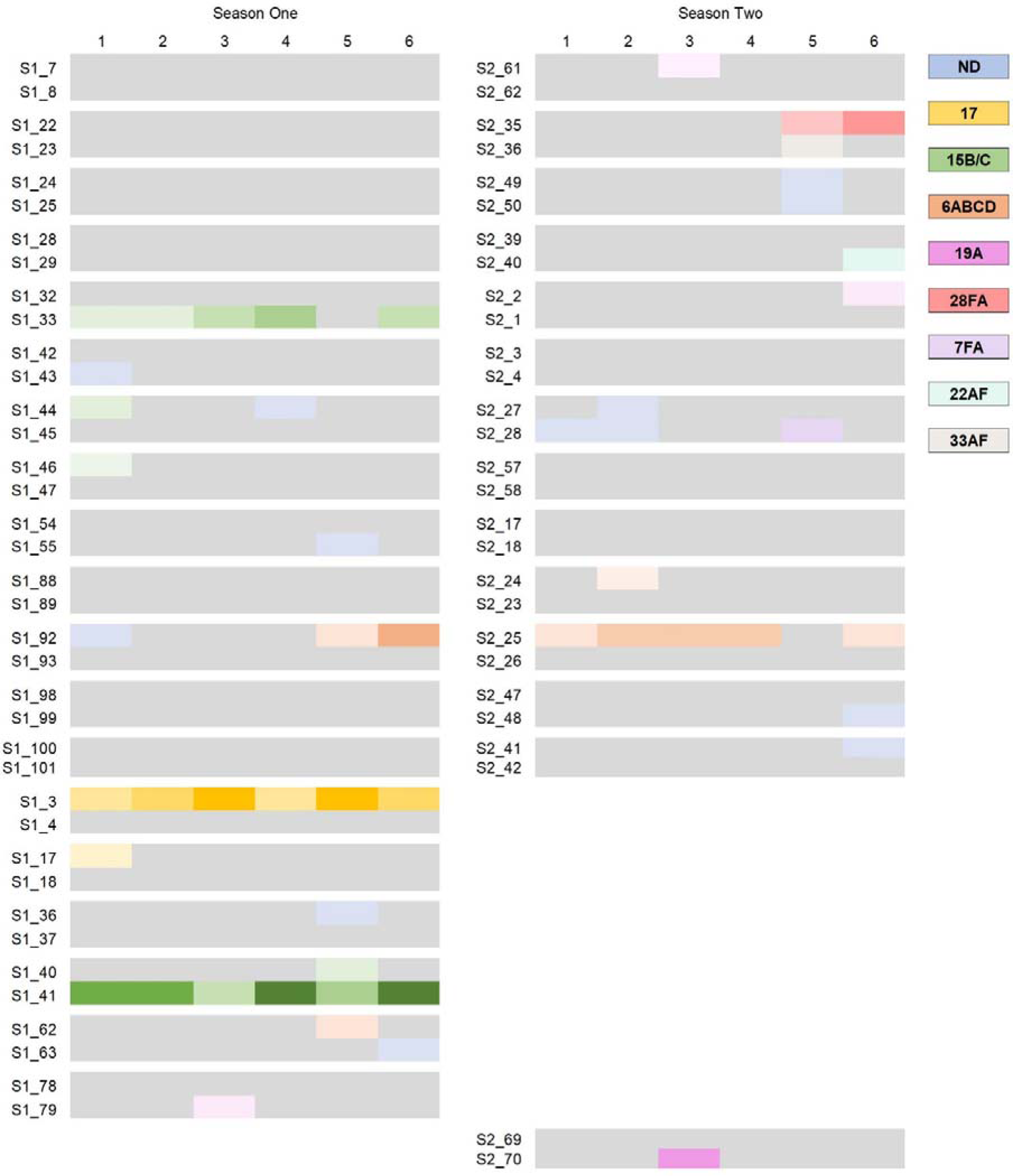
Heatmap of sample positivity for pneumococcal serotype across seasons, when tested using multiplex qPCR assays (See Supplementary Information). Darker colors indicate lower Ct values (higher concentration of serotype-specific signal). Gray boxes indicate sampling moments for which the individual tested negative for pneumococcal gene, *piaB*. Each row represents an individual, each column a time point. Individuals in the same household are grouped together. Results for study participants and households that participated in both study seasons are shown side-by-side and with the corresponding study ID. Households with results only in one column participated only in that one study season. ND: For 13 sampling moments from 12 individuals, we could not resolve a serotype due to the incomplete serotype coverage of our qPCR assays.

In line with other reports,^1,32–34^ testing *piaB*-negative DNA samples demonstrated that many serotyping assays were subject to confounding by non-pneumococcus Streptococci (See Supplementary Information). We observed high rates of false positivity with the assays targeting serotypes 9V/A,^22^ 12FAB/44/46,^35^ 17,^19^ 20,^22^ 21,^22^ 23FAB,^19^ 28F/A,^19^ and 34^22^ (much lower Ct values as compared to *piaB* or signal detected in *piaB-*negative DNA templates) and as such, all results from these assays were excluded. Interestingly, two individuals from the same household (#28 and #29), tested positive for both *lytA* and serotype 21^22^ (but negative for *piaB* and SP2020) at all 6 sampling moments. Moreover, two samples from S2 #35 tested positive for serotype 28F/A with a Ct value concordant with their *piaB* and *lytA* Ct values (within 2 Ct), yet we cannot be sure whether this represents genuine detection of a serotype 28F/A pneumococcus or a *Streptococcus* spp. with homologous genes present. We were able to replace the assays targeting serotype 9V/A,^19^ 17,^22^ and 28F/A,^36^ though 9V/A proved only marginally more specific so results from this assay were also excluded.

For 13 sampling moments from 12 individuals, we could not resolve a serotype due to the incomplete serotype coverage of our qPCR assays.

### Point prevalence was higher among those with contact with children

The point prevalence of pneumococcal carriage was substantially higher among individuals who reported contact with children as compared to those who had no recent contact with children (10.0% vs 1.6%, respectively; Table 2). Participants who reported recent contact with <5-year-olds and 5-9-year-olds had point prevalences of 14.8% and 14.1%, respectively; those reporting contact with children >10 years had a prevalence of 8.3%. While the numbers were sparse, further subdividing the <5 year-old population found high prevalence among those reporting contact with children <12m (13.8%), 12-23 months (10.7%) and 24-59 months (17.3%). While the numbers were small, those who had contact with children daily or every few days had the highest prevalence (15.7% and 14%, respectively). Those who had contact once or twice a month or no contact had lower prevalence (4.5% and 1.8% respectively: RR: 0.57, 95%CI: 0.15, 2.25; and RR: 0.33, 95%CI: 0.09, 1.18, respectively, compared with daily contact in a multivariate analysis).

**Table 2:** Percent of samples positive for pneumococcus by contact with children.

|  | n, samples tested | n, *piaB* positive | %, *piaB* positive |
| --- | --- | --- | --- |
| **OVERALL** | **1088** | **52** | **4.8** |
| **Contact with children: Any** |  |  |  |
| No | 577 | 14 | 2.4 |
| Yes | 404 | 42 | 10.4 |
| Missing/No response | 110 | 5 | 4.5 |
| **Contact with children: <5 years** |  |  |  |
| No | 781 | 27 | 3.5 |
| Yes | 196 | 29 | 14.8 |
| Missing/No response | 114 | 5 | 4.4 |
| **Contact with children: <12 months** |  |  |  |
| No | 890 | 44 | 4.9 |
| Yes | 87 | 12 | 13.8 |
| Missing/No response | 114 | 5 | 4.4 |
| **Contact with children: 12-23 months** |  |  |  |
| No | 901 | 48 | 5.3 |
| Yes | 76 | 8 | 10.5 |
| Missing/No response | 114 | 5 | 4.4 |
| **Contact with children: 24-59 months** |  |  |  |
| No | 876 | 38 | 4.3 |
| Yes | 101 | 18 | 17.8 |
| Missing/No response | 114 | 5 | 4.4 |
| **Contact with children: 5-10 years** |  |  |  |
| No | 793 | 30 | 3.8 |
| Yes | 184 | 26 | 14.1 |
| Missing/No response | 114 | 5 | 4.4 |
| **Contact with children: >10 years** |  |  |  |
| No | 809 | 42 | 5.2 |
| Yes | 168 | 14 | 8.3 |
| Missing/No response | 168 | 14 | 8.3 |
| **Intensity of contact: per day** |  |  |  |
| <4 hours (morning or afternoon) | 252 | 30 | 11.9 |
| 4-8 hours (full day) | 107 | 9 | 8.4 |
| 8+ hours (day care, overnight) | 31 | 2 | 6.5 |
| No contact/Missing/No response | 701 | 20 | 2.9 |
| **Intensity of contact: frequency** |  |  |  |
| Daily | 51 | 8 | 15.7 |
| Every few days | 172 | 23 | 13.4 |
| Once or twice a month | 158 | 9 | 5.7 |
| No contact/Missing/No response | 710 | 21 | 3.0 |

### Acquisition and clearance rates of pneumococcal colonization

The repeated sampling scheme used in this study allowed us to estimate characteristics of colonization episodes including the acquisition rate, duration of colonization, and risk factors for acquisition and clearance. The average time to acquire pneumococcus was 358 days (230 days, 556 days), indicating roughly one detected colonization episode per year. The mean duration of detectable colonization was 17.7 days (95%CI: 11.4, 27.3 days). Acquisition and duration did not vary by sex. Recent contact with children aged <10 years was associated with a significant increase in acquisition rate (hazard ratio: 3.0, 95%CI: 1.3, 7.2). Likewise, those with contact with children daily or every few days had a higher risk of acquisition than those without contact with children (hazard ratio: 6.0, 95%CI 2.3, 15.2).

## DISCUSSION

There are ongoing policy discussions about the use of PCVs in the adult population. Much of this discourse focuses on whether vaccinating adults is important in populations where children are vaccinated at high rates. Additionally, as higher-valency adult-specific PCVs are licensed and introduced into communities around the world, it is important to have baseline data of serotype colonization in children and adults and an understanding of potential pneumococcal transmission among adults and from adults to children. In this study, we found that among community-dwelling older adults, pneumococcal carriage prevalence was high. While there were households in which an individual was positive for pneumococcus across numerous sampling moments and instances where both adults in the household carried pneumococcus around the same time, there was no clear evidence of adult-adult transmission in this relatively small community-based study. Rather, carriage was highest among those who had frequent contact with young children. This suggests that the main benefit of adult PCV immunization is to directly protect adults who are exposed to children, who still carry and transmit some vaccine-type pneumococci despite successful pediatric national immunization programs. By contrast, adult PCV immunization may not have a major impact on onward transmission to other adults; whether adult to child transmission occurs was not evaluated in our study.

With the study period coinciding with the COVID-19 pandemic, we were able to explore risk factors for pneumococcal carriage in a period when strict transmission mitigation measures were in place and eased over time. Other than continued contact with families, study participants reported few activities outside of the home, adhering to the social distancing recommendations in place.^6^ This study setting allowed greater resolution into household transmission, removing many of the possible external sources that could be expected from social activities in the absence of social distancing. We found that carriage rates remained consistent across both study seasons, despite a return to community activities in the second season and an increased circulation of respiratory viruses in the local community. Other studies have reported a persistence of pneumococcal carriage in children across the COVID-19 pandemic.^37,38^ In contrast to our findings, carriage among older adults in Denmark was reported to decline.^39^ Interestingly, it was also reported that in Denmark, distancing recommendations, including the distancing from children, were strictly adhered to,^40^ unlike in our population.^6^ A limitation of our study; however, was that despite enrolling from a diverse population, or study participants were skewed towards white individuals with higher education, meaning that results may not be representative across the entire greater New Haven community.^41^ Additionally, with a third of the study participants from season one re-enrolling into season two, this further impacted the representation of the wider community.

Across the entire study, it was contact with pre-school and young school-aged children that most influenced acquisition of pneumococcus. While there were several instances where both household members were colonized at the same time or at sequential visits, these individuals also typically had contact with children. In a multivariate regression analysis, it was contact with 24–59-month-old children that was associated with increased odds of carrying pneumococcus, a finding consistent with independent modeling work in other populations.^4,5^ The frequency and intensity of contact also mattered. Higher carriage prevalence was associated with those who reported daily contact (15.7%) or contact every few days (14.0%) as compared to those who reported to have contact with children only once or twice a month (4.5%), or no contact (1.8%).

The current gold standard recommendations for the detection of pneumococcus^8^ are insensitive when applied for carriage detection in adults.^1,32,34^ As such, more sensitive methods of pneumococcal carriage surveillance must be established and appropriately applied.^42^ Saliva, when tested using qPCR, facilitates more sensitive and thus informative detection of pneumococcus in older adults.^32,43^ Therefore, in the current study, we tested saliva samples collected over two study seasons and confirmed higher carriage rates than typically reported when culture-based methods are applied to nasopharyngeal swabs. We also explored several methods for serotyping the samples. We primarily used qPCR, adapting assays that had been described elsewhere (see Supplementary Table 1) into a multiplex format. While we obtained some unambiguous findings when we focused on subjects with higher density of pneumococcal colonization (Ct<35) and were able to isolate and confirm the serotype present using traditional culture-based methods in samples with higher abundance, we were unable to resolve a primary serotype for all samples, particularly those at a lower abundance. This relatively low abundance of pneumococcus in positive samples also made it difficult to isolate pneumococcus by culture. Moreover, we observed a high prevalence of non-pneumococcal Streptococci in our samples as evident through high rates of positivity in the *lytA* qPCR assay (see Supplementary Figure 2), while negative for *piaB.*^31^ These Streptococci can have capsules that are identical to pneumococcal capsules in their structure, and whose capsular biosynthesis genes are nearly identical to pneumococcal genes.^44^ Therefore, all qPCR assays were tested on a set of samples that were *lytA* positive and *piaB* negative (indicating presence of *Streptococcus* spp., but not pneumococcus) to determine the background rate of detection. As such, we detected a high degree of non-specificity in many of the serotyping assays that have been published by others. Further validation and optimization work is required to establish more robust and reliable methods for the serotyping of pneumococci present in polymicrobial samples. Despite the improved sensitivity of detection of carriage with these methods, it is still possible that we have underestimated overall carriage prevalence because we did not sample the entire upper airway or beyond the surface of the upper airway.

With next generation PCVs being introduced into older adult populations around the world, it is crucial to establish baseline data on serotype colonization in adults as well as children, and to understand pneumococcal transmission pathways to and among adults. While a main goal of the current study was to quantify the effect of having a positive household contact on transmission, the number of households where both individuals were positive was extremely low, and it was contact with children that was most highly associated with colonization in this population. In addition to diagnostic testing differences, differing rates of contact with children could also help explain the differing pneumococcal colonization prevalences reported from different populations of older adults around the world. In other contexts, such as nursing homes, more adult-adult transmission likely occurs as this would explain nursing home based pneumococcal disease outbreaks.^45^ To adequately investigate this however, the establishment and appropriate application of more sensitive methods for pneumococcal carriage surveillance are imperative. Despite some limitations, the results of our study contribute to our understanding of pneumococcal carriage and can inform future policy decisions and preventive strategies aimed at reducing pneumococcal disease burden in children and adults.

## Data Availability

All data produced in the present study are available upon reasonable request to the authors

## ACKNOWLEDGEMENTS

We thank the study participants for their time and dedication to our study. We thank all members of the research team of the Wyllie and Weinberger Laboratories at the Yale School of Public Health and the team at the Yale Center for Clinical Investigation for their dedication and work which made this project possible. In particular, we would like to thank Harold Kennedy for assistance with sample transport and Molly McLaughlin for assistance with participant consent.

## ROLE OF THE FUNDER

This study was conducted as a collaboration between Yale School of Public Health and Pfizer. Yale School of Public Health is the study sponsor. The study protocol was designed by the Yale researchers in consultation with Pfizer. The decision to publish was made by the Yale researchers in consultation with Pfizer; all authors agree with the decision to publish and with the results of the study.

## AUTHOR’S CONTRIBUTIONS

ALW, AA and DMW conceived the study. ALW, RAP, AA, BDG and DMW designed the study protocol. ALW, DY-C, and DMW managed the study. DY-C and HK collected the data. DY-C, SM, and AY were responsible for sample receipt, processing and testing. ALW and DMW performed the analyses and interpreted the data. ALW and DMW drafted the manuscript. All authors amended and commented on the final manuscript.

## DISCLOSURES

ALW has received consulting and/or advisory board fees from Pfizer, Merck, Diasorin, PPS Health, Primary Health, Co-Diagnostics, and Global Diagnostic Systems for work unrelated to this project, and is Principal Investigator on research grants from Pfizer, Merck, NIH RADx UP and SalivaDirect, Inc. to Yale University and from NIH RADx, Balvi.io and Shield T3 to SalivaDirect, Inc.. DMW has received consulting fees from Pfizer, Merck, GSK, and Matrivax for work unrelated to this project and is Principal Investigator on research grants and contracts with Pfizer and Merck to Yale University.

## Notes

### Author Declarations

This study was approved by the Institutional Review Board at Yale School of Medicine (Protocol ID #2000026100). Demographic data and samples were collected after the study participant had acknowledged that they had understood the study protocol and provided digitally signed-informed consent, collected in the Research Electronic Data Capture (REDCap) electronic data capture tools hosted at Yale University. All participant information and samples collected were assigned anonymized study identifiers.

