## Supplementary for "Contact with young children is a major risk factor for pneumococcal colonization in older adults"

Anne L. Wyllie^1^*, Devyn Yolda-Carr^1^, Maikel S. Hislop^1^, Sidiya Mbodj^1^, Loren Wurst^1^, Pari Waghela^1^, Ronika Alexander-Parrish^2^, Adriano Arguedas^2^, Bradford D. Gessner^2^, Daniel M. Weinberger^1^*

^1^Department of Epidemiology of Microbial Diseases, Yale School of Public Health, New Haven, CT 06510, USA; ^2^Pfizer Inc.

*These authors contributed equally to the study

**Corresponding author:**

Anne Wyllie, PhD

Yale School of Public Health

LEPH 823

60 College St

New Haven

CT 06510

USA

**Running title**: Pneumococcal carriage in older adults

**Keywords:** pneumococcus, saliva, surveillance, carriage, transmission

| **Table S1: Percent of samples positive for pneumococcus by demographic or health status** | | | |
| --- | --- | --- | --- |
|  | **n, samples tested** | **n, *piaB* positive** | **%, *piaB* positive** |
| **OVERALL** | **1088** | **52** | **4.78** |
| **Season** |  |  |  |
| S1 | 567 | 31 | 5.5 |
| S2 | 521 | 21 | 4.0 |
| **Gender** |  |  |  |
| Female | 561 | 24 | 4.3 |
| Male | 527 | 28 | 5.3 |
| **Symptoms: nasal congestion** |  |  |  |
| No | 877 | 47 | 5.4 |
| Yes | 110 | 3 | 2.7 |
| Missing | 101 | 2 | 2.0 |
| **Symptoms: runny nose** |  |  |  |
| No | 888 | 45 | 5.1 |
| Yes | 99 | 5 | 5.1 |
| Missing | 101 | 2 | 2.0 |
| **Recent antibiotics** |  |  |  |
| No | 766 | 39 | 5.1 |
| Yes | 65 | 2 | 3.1 |
| Missing | 257 | 11 | 4.3 |
| **Recent flu shot** |  |  |  |
| No | 32 | 1 | 3.1 |
| Yes | 895 | 42 | 4.7 |
| Missing | 161 | 9 | 5.6 |
| **Diabetes** |  |  |  |
| No | 920 | 36 | 3.9 |
| Yes | 62 | 7 | 11.3 |
| Missing | 106 | 9 | 8.5 |
| **Asthma** |  |  |  |
| No | 910 | 43 | 4.7 |
| Yes | 36 | 1 | 2.8 |
| Missing | 142 | 8 | 5.6 |
| **Education level** |  |  |  |
| High School | 98 | 3 | 3.1 |
| Undergraduate (Bachelor or Associate) | 418 | 12 | 2.9 |
| Graduate | 431 | 21 | 4.9 |
| Unknown | 141 | 16 | 11.3 |

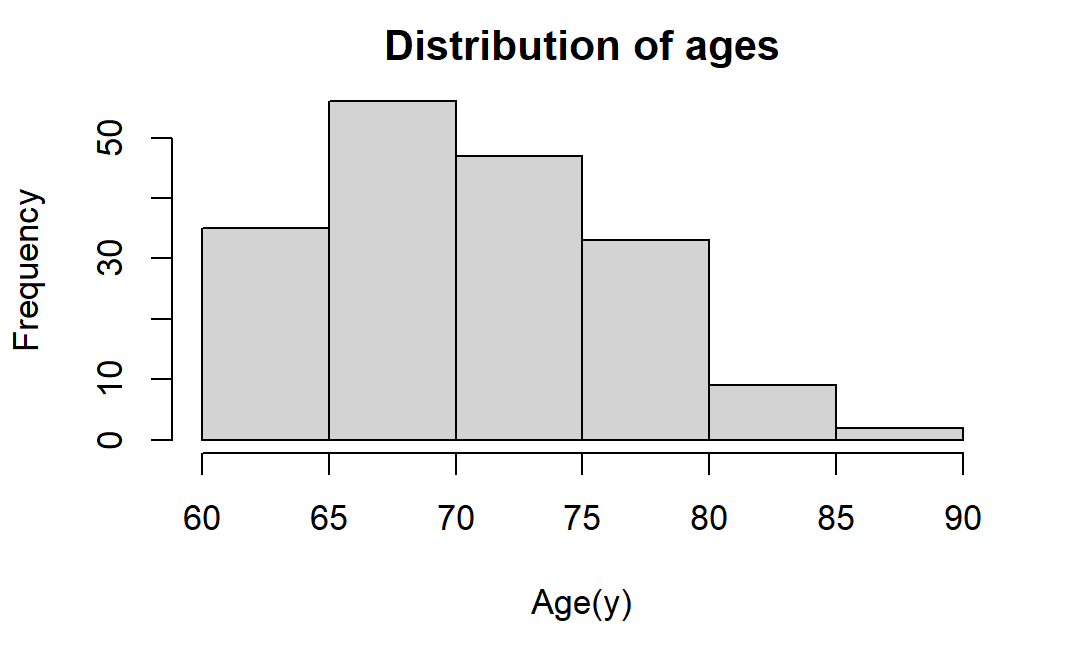

**Supplementary Figure 1.** Distribution of ages (years) of enrolled participants.

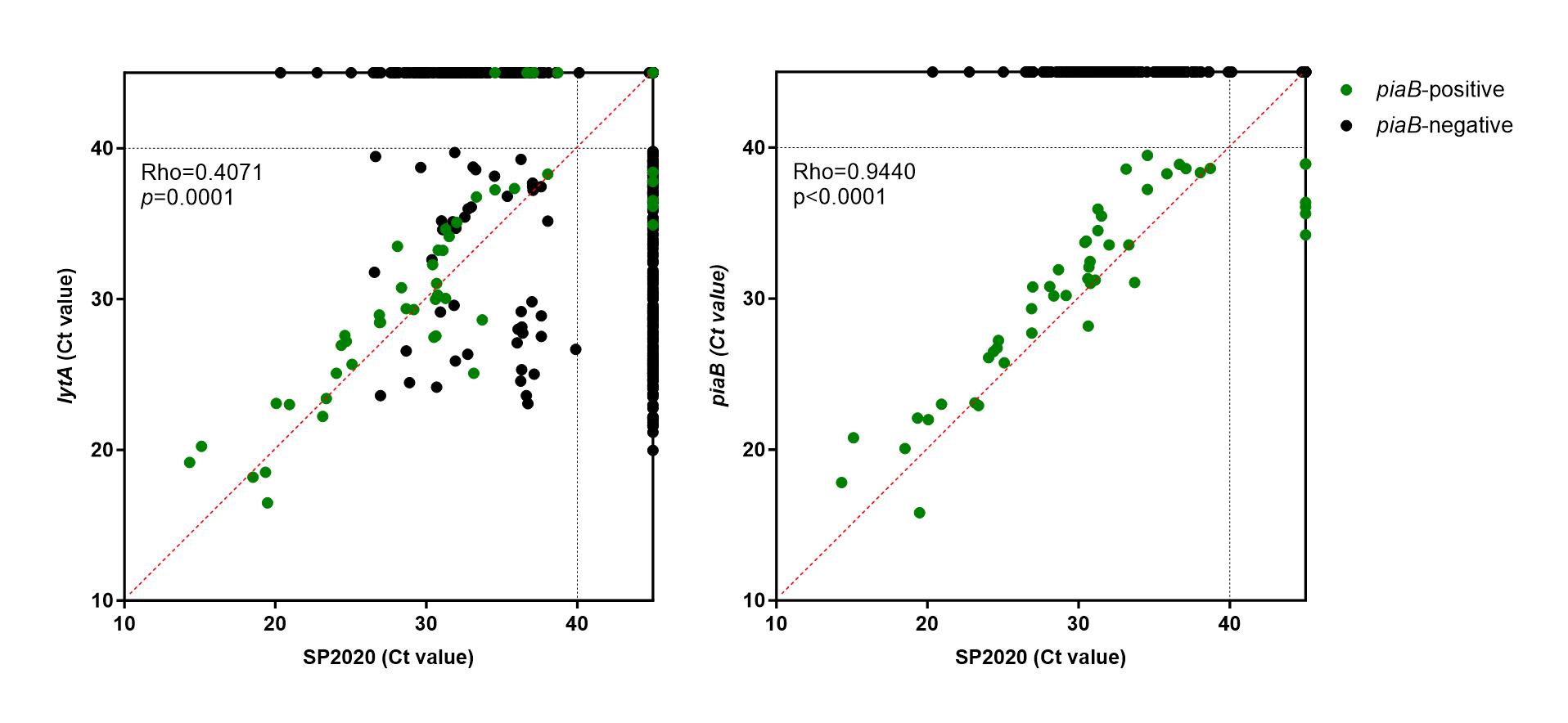

**Supplementary Figure 2.** Concordance between SP2020 and (**A**) *lytA* and (**B**) *piaB* qPCR values. Ct values <40 are considered positive. Spearman's rank correlation coefficient (rho) and the associated *p*-value (*p*) for samples generating Ct values <40 for both targets are shown. Samples shown in green tested positive for *piaB* and were interpreted as positive for pneumococcus.

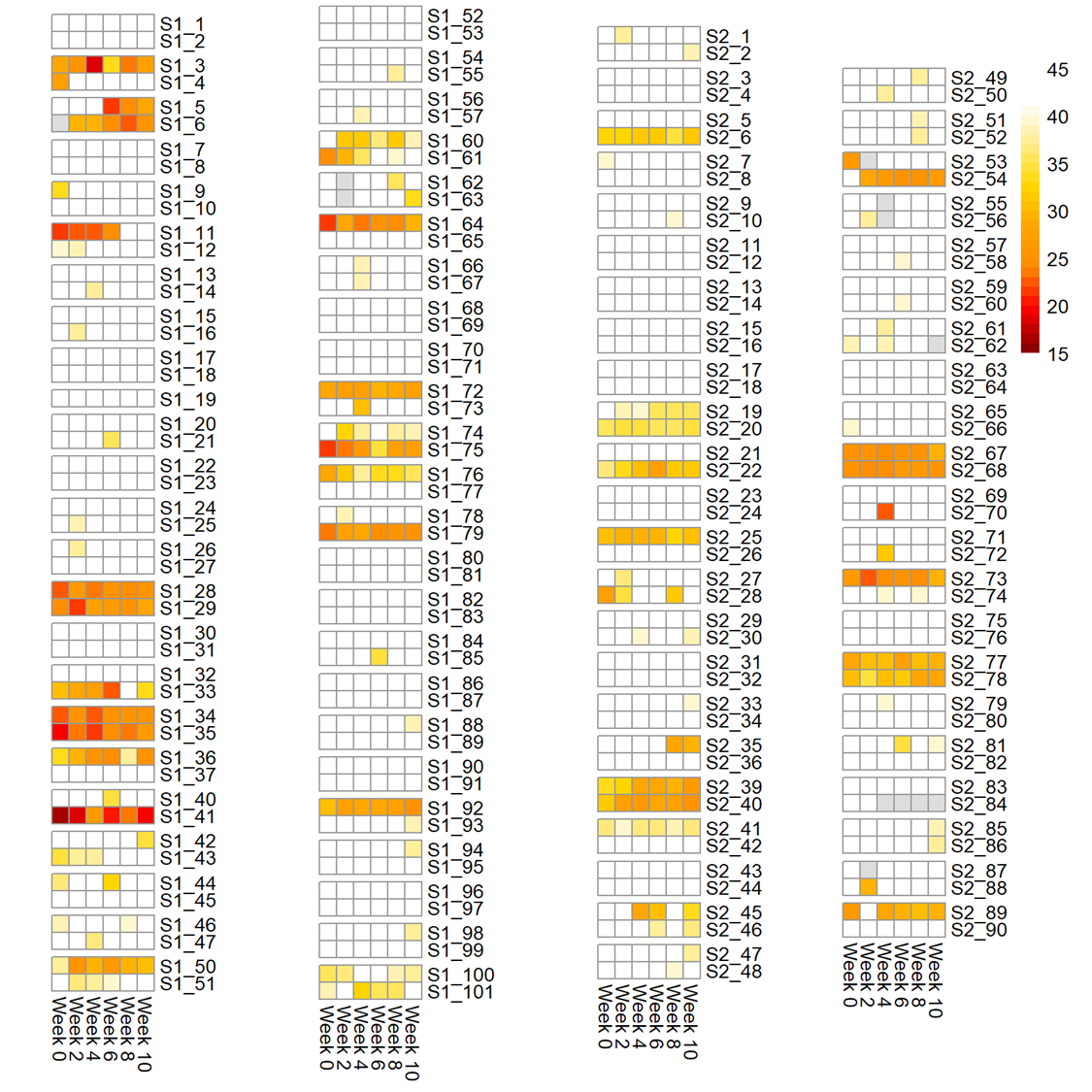

**Supplementary Figure 3.** Heatmap of sample positivity for pneumococcus gene, *lytA*. Overall, 27.7% of samples tested positive for *lytA* in season one (S1) and 24.8% samples tested positive in season two (S2). Darker colors indicate lower Ct values (higher concentration of *lytA*). Gray boxes indicate the sample was not tested or missing. Each row represents an individual, each column a time point. Individuals in the same household are grouped together.
